# Cross-national comparison of the relationship between working hours and employment status and sleep duration and quality among Australia, Germany, Japan and the United Kingdom. A research protocol

**DOI:** 10.1101/2024.06.05.24308486

**Authors:** Ya Guo, Senhu Wang, Rong Fu, Jacques Wels

## Abstract

**Background:** There is a significant gap in sleep duration across countries with 56 percent of the Japanese population sleeps less than seven hours per day against around 30 percent in the United Kingdom (UK), Germany, and Australia. Similarly, labour market characteristics differ across these countries, with average working hours being higher in Australia and Japan compared to the UK and Germany, but with a significant number of contract and part-time workers. This research aims to address how employment status and working time associate with sleep time and sleep quality across Japan, the UK, Germany, and Australia.

**Methods:** We use and harmonize four representative panel datasets, Understanding Society in the UK, the Japan/Keio Longitudinal Panel Survey in Japan, the German Socio-Economic Panel, and the Household, Income and Labour Dynamics in Australia. We use parallel analyses and focus on years 2009 to 2021 and include all respondents aged 20 to 69. We use fixed and mixed effects model using sleep time (log) as a linear outcome and self-reported sleep quality, trouble falling asleep and loss of sleep over worry as binary outcomes. Exposures include the employment status (model 1), working time (model 2) and the interaction between employment security and working time (model 3). We control for socio-demographic and socio-economic layers of adjustment and analyses are stratified by age-group, gender and whether respondents work in professional occupations.

**Results:** Expected results include descriptive statistics on sleep time and quality, employment status distribution and working time among four countries and details by gender. Estimates from fixed and mixed effects are compared using a Hausman test. Coefficients are shown for sleep time and other sleep quality measurements are in odds ratios with 95%CI.

**Conclusion:** Using parallel analyses on four panel datasets from Japan, the UK, Germany, and Australia, the study will address to what extent sleep duration and quality vary and sub-groups and whether employment status and working time patterns contribute to explain sleep differences across both countries.

**Funding:** JW receives funding from the following sources: the European Research Council (ERC) and the Belgian National Scientific Fund (FNRS).

**Conflicts of interests:** The authors report no conflict of interest. JW is a member of the Belgian Health Data Agency (HAD) user committee.

**Data availability:** Access to the Japan Household Panel Survey micro-data is available upon request via the Keio University (Japan) research portal: https://www.pdrc.keio.ac.jp/en/paneldata/datasets/jhpskhps/

Access to Understanding Society (UKHLS) micro-data is available upon request via the UK data archive: https://www.data-archive.ac.uk/

Access to German Socio-Economic Panel (SOEP) micro-data is available upon request via: https://www.eui.eu/Research/Library/ResearchGuides/Economics/Statistics/MicroDataSet Access to the Household, Income and Labour Dynamics (HILDA) micro-data is available upon request via: https://dataverse.ada.edu.au/dataverse/DSSLongitudinalStudies

## Background

Average sleep duration significantly varies across countries. For instance, 56 percent of the Japanese population sleeps less than seven hours per day against 35 percent in the United Kingdom, 31 percent in Australia, and 30 percent in Germany ^1^. Several country features explain such a gap including demographic composition and employment characteristics.

In Japan, sleep duration has been declining since the 1960s ^2^. In 2014, “good sleep” became a policy priority with specific sleep guidelines published for different generations ^3^ although mainly focusing on individuals’ behaviours and not targeting the social mechanisms leading to poor sleep quality and short sleep duration among which work and employment are usually seen as detrimental ^4,5^ . As a matter of fact, despite regulations passed over the past decades to prevent long working hours, working time remains high in Japan ^6^ and the labour market is fragmented with many women – particularly among the oldest generations – remaining out of the labour market (as housekeepers) ^7^ and older workers sometimes downgraded to specific employment statuses (contract work) ^8,9^. High working time and a fragmented labour market come together with profound demographic changes. By 2050, it is estimated that the percentage of people aged 65 and over will reach 37.5 percent of the population. It was 4.9 percent in 1950 and 17.8 percent in 2000. By contrast, it will reach 28.9 in Europe in 2050 and was 7.9 and 19.1 in 1950 and 2000 ^10^. As ageing explains alteration in normal sleep pattern and comes with lower and less regular sleeping time ^11^ and sleep disturbance^12^, Japan faces multiple challenges to improve population sleep.

The relationship between employment and sleep is different in the United Kingdom. By comparison with Japan, the UK population now sleeps more than they used to in the 1970s as they go to bed earlier and wake up later. This is true also for the employed population for which an increase in sleeping time was observed over the same period ^13^. Nevertheless, differences exist when looking a micro data. Whilst working time is lower, employment statuses are less fragmented and the ageing of the population is less pronounced in the UK, employment remains a major driver of sleep discrepancies across the British population. First, the relationship between working hours and sleep duration is also present in the United Kingdom where there is an inverse relationship between working hours and sleep duration that is stronger among the male population ^14^. Second, socio-economic differences – that are more pronounced in the UK – although not having a clear impact on sleep quantity ^15^ are associated with poor sleep quality as people from socio-economically deprived groups are at increased risk of poor sleep ^16^.

Additionally, studying the relationship between employment and sleep in Germany is crucial for understanding how labour dynamics affect individual well-being in a highly industrialized and economically significant country. According to the 2017 Health Report of the German Statutory Health Insurance (DAK), approximately one-third of employed German adults aged 35 to 65 report experiencing insomnia at least three times per week. Compared to 2010, sleep problems and disorders have increased by approximately 60%^17^. Germany’s strong emphasis on work-life balance, robust labour laws, and cultural attitudes towards health and wellness make it an ideal setting for such research^18^. Notably, Germans generally prioritize sleep as an essential component of health, with average sleep durations typically aligning with recommended guidelines of 7-9 hours per night. However, variations exist across different demographics and employment sectors, with factors such as job stress, shift work, and long working hours often contributing to sleep deprivation or poor sleep quality among certain groups. This investigation provides valuable insights into how employment conditions influence these variations in sleep patterns and overall health. Additionally, as Germany undergoes demographic changes and technological advancements, understanding these interactions becomes essential for developing effective policies to promote both worker productivity and public health.

Studying the relationship between employment and sleep in Australia is important due to its unique socio-economic landscape and labour market characteristics. Australian adults report sleeping on average, 7–8 hours a day^18^, which is within the recommendations outlined in Australia’s Sleep Health Foundation guidelines. Despite this, 66% adults report at least one sleep problem and almost half of all adults report at least two sleep related problems^20^. Factors such as long working hours, shift work, and high job demands contribute to sleep deprivation among certain groups. Additionally, the prevalence of digital devices and lifestyle choices also impact sleep quality and duration. Australia has a diverse workforce with significant representation of both urban and remote workers, and the labour market is influenced by these factors. The growing gig economy and flexible work arrangements present new challenges and opportunities for understanding sleep patterns among workers^20^. The impact of these employment conditions on sleep is further compounded by cultural attitudes towards work and health, making it essential to examine how these dynamics affect sleep duration and quality. Insights from this research can inform targeted interventions and policies to enhance worker well-being, productivity, and public health, while also addressing the unique needs of Australia’s diverse workforce.

To better understand differences between Japan, the UK, Germany, and Australia, one should account for the interplay between employment and working time. Working time is a complex variable when it comes to cross-national comparison. According to the OECD, average hours actually worked per year per worker were 1,707 in Australia,1607 in Japan, 1532 in the UK, and 1341 in Germany in 2022 ^21^. However, average hours worked do not translate sub-population differences. In Japan, long working hours remain the norm among the male working population, contributing to shape employed women’s career path and leading to high rates of part-time work within the female population ^22^. This long working hours pattern explains sleeping time differences across the Japanese population. Comparing white collar Japanese employees with the rest of the population, a cross-sectional study ^23^ has shown a higher prevalence of poor sleep quality (based on the Pittsburgh Sleep Quality Index (PSQI)) – between 30 and 45 percent – among the former. Significant factors associated with poor sleep included stress, job dissatisfaction, being unmarried, lower education, younger age and poor sleep quality was associated with absenteeism, poor health, work and relationship problems and workplace accidents. Mafune and Yokoya ^24^ have shown that workers with over 100 hours of overtime experiencing less than 6 hours of sleep, late dinners, and increased dining out. Night shift workers also reported more frequent awakenings during sleep. The conclusion highlighted that around 30% of the surveyed temporary workers were at risk of overwork-related health issues, including insufficient sleep, late meals, and mental health symptoms, suggesting a need for regulations to prevent excessive overtime requests. Early start of the working day is associated with lower sleep duration, sleep problems, and fatigue ^25^. Working time is also associated with sleep duration, with those working less that 8 hours a day sleeping more than those working more than 8 hours a day ^26^.

However, non-employed people are also at risk. It was evidenced that unemployment (i.e., those looking for a job) and non-employment are associated with high insomnia-related symptoms prevalence ^27^, particularly among the male population and to a lesser extent among the female population ^28^. A substantial amount of research was produced on the relationship between transition from work to retirement and sleep quality and duration. It was shown that retirement is not only associated with short-term reductions in sleep difficulties but also increase in sleep duration over 1 to 2 years ^29,30^. Results are similar using panel data from France ^31^. Using longer follow-up longitudinal data, it was demonstrated that these positive effects last over time for non-restorative sleep, premature awakening and restless sleep ^32^ with potential greater effects on female as well as greater effects on those retiring from part-time jobs ^33^. Both employment and non-employment may affect sleep duration and quality through different mechanisms but the amount of evidence is limited when it comes to addressing short sleep and poor sleep quality across different statuses ^34^.

The relationship between work and employment and sleep is of interest in such a context as low sleep duration and poor sleep quality both have detrimental effects with sleep disturbances associated with depressive symptoms among the older population ^35^ and low (<6 h) and high sleeping time (>9h) associated with higher mortality risks ^36^. However, national contexts also explain both sleep patterns across populations and the way employment and working time affect sleep. Only a few studies have addressed such a relationship using longitudinal cross-national data. Comparison panel data on the population aged 20 to 69 among the UK, Germany, Australia, and Japan, this study aims to address the following three research questions: (1) *Do sleep duration and quality vary among Japan, the United Kingdom, Germany, and Australia, and do differences occur within population sub-groups including the employed or female populations?* ; (2) *Are some specific employment status associated with lower sleep duration and poorer sleep quality after controlling for individual socio-economic characteristics?* ; (3) *Are the associations between working time and sleep duration and quality similar across countries as well as by gender and type of occupation?*.

## Data and methods

### Understanding Society (USoc) and the Japan Household Panel Survey (JHPS)

The <u>Japan Household Panel Survey (JHPS)</u> includes two longitudinal datasets: the Keio Household Panel Survey (KHPS) and the Japan Household Panel Survey (JHPS). Both datasets can be combined. The KHPS data collection started in 2004 (with a baseline sample size of N=4,005 respondents) and yearly data have been collected since then. Two refreshment samples were added to the original 2004 cohort in 2007 (N=1,419) and 2012 (N=1,012). The JHPS data collection started in 2009 (N=4,022) and a new cohort was added in 2019 (N=2,203). Sample section in KHPS and JHPS is based on the same methodology. Respondents are selected through a two-sage stratified random sampling. In the first stage, Japan is stratified into 24 levels based on regional and city classification and the sample is distributed across 24 levels based on resident register population. In the second stage, sample is selected based on gender and age group. The population target is similar but does not overlap within both datasets but KHPS includes respondents aged 20 to 69 whilst JHPS people includes respondents aged 20 and above. When combined, the population should be restricted to respondents aged 20 to 69. Questionnaires were homogenised in 2009 and whilst some core variables were similar in pre-2009 KHPS, correspondence might be an issue when using pre-2009 data. KHPS and JHPS provide both longitudinal and cross-sectional weights based on the cohort used for analyses. Current data are available until 2021.

<u>Understanding Society (USoc)</u>, the UK Household Longitudinal Study (UKHLS), currently contains 13 waves collected 2009-10 (wave 1), 2010-11 (wave 2), 2011-12 (wave 3), 2012-13 (wave 4), 2013-14 (wave 5), 2014-15 (wave 6), 2015-16 (wave 7), 2016-17 (wave 8), 2017-18 (wave 9), 2018-19 (wave 10), 2019-20 (wave 11), 2020-21 (wave 12) and 2021-22 (wave 13). UKHLS has a clustered and stratified, probability sample of 24,000 households living in Great Britain in wave 1 and a random sample of 2,000 household in Northern Ireland. The sample is 54,559 in 2010-11, 44,071 in 2012-13; 44,396 in 2014-15; 38,782 in 2016-17; 33,588 in 2018-19; and 28,520 in 2020-21.

<u>The German Socio-Economic Panel (SOEP)</u> is a longitudinal survey of approximately 15,000 private households in the Federal Republic of Germany from 1984 to 2021 and the eastern German länder from 1990 to 2021 (release 2023). These waves, spanning over several decades, provide a comprehensive and longitudinal view of socio-economic trends and life trajectories within the German population.

<u>The Household, Income and Labour Dynamics (HILDA)</u> survey is a household-based panel study that collects information on many aspects of life in Australia, including household and family relationships, income and employment, and health and education, including working time mismatch, work-life balance, employment status, and other important demographic and socioeconomic characteristics. The study collected data annually through interviews with all people over 15 years old in each household. In wave one, the study collected data from 7,682 households (13969 individuals). The average response rate exceeds 60%.

For comparability purpose, both datasets are restricted to the population aged **20 to 69** for years **2009 to 2021**.

### Outcomes: Sleep duration and quality

The four datasets include variables on sleep duration and quality with slight differences in definitions, modalities and time-point collection. Information of variable availability across waves within the different datasets is shown in <u>supplementary file S1</u> whilst information on variables’ questions and response modalities are shown in <u>supplementary file S2</u>.

- *Sleep duration* is calculated as the average number of hours respondents report sleeping by night. Information on sleeping time in USoc is available at waves 2009, 2012, 2015, 2018 and 2021, at waves from 2008-2021, and in all waves for JHPS and HILDA. USoc does not specific whether this sleeping time relates to weekdays or weekends whilst JHPS specifically asks about sleeping hours during weekdays for all waves. JHPS also include a question on sleeping hours during weekends in waves 2011 to 2021. The main analyses were made using the average sleeping time in USoc and the average weekdays sleeping time in JHPS but we include an additional analysis for JHPS and SOEP where the average of weekdays and weekend sleeping times is calculated (<u>sensitivity analysis 1</u>). We calculate the ***natural logarithm*** of the variable and use it as *numeric*.
- *<u>Troubles falling asleep</u>* refers to the porosities of not being able to fall asleep within 30 minutes over the last months in Usoc (from 1. Not at all to 4. Much more than usual) whilst JHPS and SOEP only ask whether respondents have trouble getting sleep (from 1. Never to 4. Often). The HIILDA asks considerate questions about troubles falling asleep. The variable is available between waves 2009 and 2013 in JHPS and in waves 2012, 2015, 2018, 2021. The variable is available between in waves 2011, 2012, 2016, and 2021 in SOEP and in waves 2013, 2015, and 2017 in HILDA. We transform the variable into a **binary outcome** using modalities 1 and 2 as the reference and 3 and 4 as “1”.
- *<u>Lost sleep over worry</u>* refers to loss of sleep calculated on a 4-items scale coded from (1) Never to (4) Often in JHPS and (1) Not at all to (4) Most of the time in Usoc. The variable is available from 2009 to 2021 in USoc as part of the General Health Questionnaire questions set and from 2014 to 2019 in JHPS. HILDA has not collected this measure. We transform the variable into a **binary outcome** using modalities 1 and 2 as the reference and 3 and 4 as “1”.
- *<u>Sleep quality</u>* refers to the answer to the following questions: “During the past month, how would you rate your sleep quality overall?” in USoc and HILDA and “How would you rate the overall quality of your sleep over the past month?” in JHPS. SOEP asks the respondents’ satisfaction with sleep. The answer modalities are coded from (1) Excellent to (4) Very bad in JHPS and from (1) Very good to (4) Very bad in USoc. The variable is available in waves 2012, 2015, 2018 and 2021 in Usoc but only in wave 2021 in JHPS. We transform the variable into a **binary outcome** using modalities 1 and 2 as the reference and 3 and 4 as “1”.
- Information on the *<u>use of sleeping medicine</u>* coded on a binary basis (yes/no) is also available in Usoc at waves 2012, 2015, 2018, 2021, and in HILDA at waves 2013, 2015, and 2017 but not in JHPS and SOEP.

Additional analyses are made on troubles falling asleep, lost sleep over worry and sleep quality using the last available modality (4: most of the time) as the modality of interest <u>as sensitivity check</u>.

### Exposures: Employment and working time

The study includes three models where exposure variables are derived from the employment status and working time.

**Model 1** focuses the <u>full-sample</u> and uses the <u>employment status</u> as the exposure variable. To ensure homogeneity across the four datasets, we have derived composite employment status variables containing the following modalities: (1) permanent employment (part-time and full-time), (2) temporary employment or contract work, (3) self-employment, (4) unemployment, (5) inactivity (<u>reference category)</u>. The employment status in JHPS, SOEP, and HILDA include whether the respondent is employed (full-time or part-time), self-employed, working under contract (i.e., contract work), inactive or unemployment. The employment status in USoc does not directly distinguishes permanent and non-permanents types of job but an additional question exist on whether the current job is permanent or temporary (leaving aside respondents’ intentions or circumstances). Note that in both variables, we are using inactivity as the reference category, principally because unemployment rates are low in Japan.

**Model 2** focuses on the <u>full-sample</u> and uses working time as the exposure variable. In USoc and JHPS, working time is the average working time worked per week including overtime and is coded using the following modalities: (1) 1 to 20 hours/week; (2) 21-34 hours/week; (3) 35-48 hours/week; (4) 48+ hours/week; (5) not in employment (reference). In SOEP and HILDA, working time is the average working time worked per week in main jobs.

**Model 3** restricts the sample to the <u>employed population</u> only (excluding unemployed, inactive and self-employed respondents) and includes an interaction term between working time (used as a categorical variable with 35-48 hours/week as the reference category) and whether the job is permanent or not (including contract work).

### Control variables and adjustment levels

The study includes several fixed and time-varying control variables that were harmonized across the four studies.

Fixed variables include gender (‘male’ is the reference), age and age-square as well as the highest level of education distinguishing those with and without a university degree (university degree is the reference category).

Time-varying variables include the marital status (non-married or married (reference)), the presence of a child or children (no or yes (reference)), the logged household net incomes, whether the respondents have caring responsibilities (no or yes (reference)), self-reported health (numeric, from 1 (poor) to 5 (very good -- reference)), whether the respondent has a longstanding illness (yes or no (reference)).

The models include several levels of adjustment. The <u>unadjusted model</u> does not control for any covariates in the fixed effects model whilst it controls for age, age-square, gender and level of education in the random effect. The <u>socio-demographic adjusted</u> model additionally controls for marital status, presence of children, logged household incomes and whether the respondents has caring responsibilities. The <u>health adjusted model</u> additionally controls for self-reported health and the presence of a longstanding illness.

### Analyses

We provide descriptive statistics on the following. First, we show the distribution of sleep duration, trouble falling asleep, lost sleep over worry, sleep quality and use of sleep medicine across the full population, across the self-employed and employed population only and across the female population only (for all waves). Second, we provide means by year of data collection for all these variables for the fully population, the self-employed and employed population and across the female population only. Third, we provide working time distribution for all waves over the full employed and self-employed population and over the female population. Fourth, we provide average working time by year of data collection for the full population and the female population. All descriptive statistics are produced using cross-sectional weight to ensure representativeness and a 95 percent confidence interval is calculated for the yearly means.

We then use parallel analysis in which each estimates are calculated separately for the four datasets to maximize survey similarities and differences. We provide estimates from both a two-way fixed effects linear model and a mixed effects linear model using respectively sleeping hours (log), trouble falling asleep (binary), lost sleep over work (binary), sleep quality (binary) and whether respondents take sleep medicine (binary) as the outcome variables. Use of sleeping medicine information is only collected in USoc and HILDA and no results are provided for JHPS and SOEP. The model is replicated twice including the employment status as exposure (model 1) and on a sample restricted to the employed population with working time as the exposure (model 2) and the interaction between working time and whether the job is permanent (model 3). We perform a Hausman test to compare fixed and mixed effect estimates (only for the fully adjusted models).

### Stratification

Models 1, 2 and 3 are stratified using the following variables:

- <u>Gender</u>: Male / female
- <u>Age group</u>: 20-39, 40-59, 59-69.

Models 2 and 3 are stratified using the following variable:

- <u>Professional and non-professional occupations</u>: USoc distinguishes professional and non-professional occupations based on the National Statistics Socio-Economic Classification (NS-SEC) including the following categories: managements & professional, intermediate, small employers & own account, lower supervisory & technical and semi-routine & routine. NS-SEC nomenclature cannot be replicated with JHPS, but several employment statuses are distinguished within the study including full-time regular employee with no title, full-time regular employee with title, full-time regular employee manager, contract employee, part-time worker, subcontracted worker, and specialized contract employee. SOEP and HILDA also collected the respondents’ current industry occupations. For harmonization, we distinguish management & professional and non-professional occupations accordingly.

Note: stratification analyses will be made on the fixed or mixed effect only depending on the results from the Hausman test in the non-stratified models.

### Weights and missing data

Descriptive statistics are produced using provided cross-sectional weights. Fixed and random effects are produced using specific longitudinal weights provided within each dataset.

### Descriptive statistics Data to plot histograms

**<u>Table 1:</u>** <u>Sleep hours distribution (percentage by sleeping hours) for the full population and for all waves together (cross-sectional weight)</u>

**<u>Table 2:</u>** <u>Sleep hours distribution (percentage by sleeping hours) for the working population only (excluding unemployed and inactive respondents) and for all waves together (cross-sectional weight)</u>

**<u>Table 3:</u>** <u>Sleep hours distribution (percentage by sleeping hours) for the female population only (excluding male) and for all waves together (cross-sectional weight)</u>

**<u>Table 4:</u>** <u>Working hours distribution (percentage by working hours) for the working population only (excluding unemployed and inactive respondents) and for all waves together (cross-sectional weight)</u>

**<u>Table 5:</u>** <u>Working hours distribution (percentage by working hours) for the female population only (excluding male respondents) and for all waves together (cross-sectional weight)</u>

### Data to plot time-series (2009-2022)

**<u>Table 6:</u>** <u>Mean sleep hours per year for the non-working population and the working population (employed and self-employed) (cross-sectional weight) including 95%CI.</u>

**<u>Table 7:</u>** <u>Mean sleep hours per year by gender (cross-sectional weight) including 95%CI.</u>

**<u>Table 8:</u>** <u>Percentage of trouble falling asleep per year for the non-working population and the working population (employed and self-employed) (cross-sectional weight) including 95%CI.</u>

**<u>Table 9:</u>** <u>Percentage of trouble falling asleep per year by gender (cross-sectional weight) including 95%CI.</u>

**<u>Table 10:</u>** <u>Percentage of lost sleep over worry per year for the non-working population and the working population (employed and self-employed) (cross-sectional weight) including 95%CI.</u>

**<u>Table 11:</u>** <u>Percentage of lost sleep over worry per year by gender (cross-sectional weight) including 95%CI.</u>

**<u>Table 12:</u>** <u>Percentage of poor sleep quality for the non-working population and the working population (employed and self-employed) (cross-sectional weight) including 95%CI.</u>

**<u>Table 13:</u>** <u>Percentage of poor sleep quality by gender (cross-sectional weight) including 95%CI.</u>

**<u>Table 14:</u>** <u>Percentage of employment status for the working population per year including (1) permanent employment (part-time and full-time), (2) temporary employment or contract work, (3) self-employment, (4) unemployment, (5) inactivity (cross-sectional weight) including 95%CI.</u>

**<u>Table 15:</u>** <u>Percentage of employment status by gender (male/female) for the working population per year including (1) permanent employment (part-time and full-time), (2) temporary employment or contract work, (3) self-employment, (4) unemployment, (5) inactivity (cross-sectional weight) including 95%CI.</u>

### Main estimates to be produced

#### MODEL 1

##### Model 1.a. Two-way fixed effects (including longitudinal weight)

***Outcomes***: Sleeping hours (numeric, log) / trouble falling asleep (binary) / lost sleep over worry (binary) ^#^ / sleep quality (binary) / take medicine to sleep (binary)*

*Notes:* ^#^ *Not for HILDA, * Only for Usoc and HILDA*

***Exposure:***

Exposure 1=Employment status (reference: inactive)

***Adjustment:***

~~~
           1.a.a.: none
           1.a.b.: + Marital status, presence of children, logged household incomes,
           caring responsibilities, professional occupation
           1.a.c.: + self-reported health, longstanding illness
~~~

**Stratification:**

~~~
           1.a.d.: Male (fully adjusted model (1.a.c.) only)
           1.a.e: Female (fully adjusted model (1.a.c.) only)
           1.a.f.: age-group 20-39 (fully adjusted model (1.a.c.) only)
           1.a.g: age-group 40-59 (fully adjusted model (1.a.c.) only)
           1.a.h: age-group 59-69 (fully adjusted model (1.a.c.) only)
~~~

#### Model 1.b. Mixed effect (including longitudinal weight)

***Outcomes***: Sleeping hours (numeric, log) / trouble falling asleep (binary) / lost sleep over worry (binary) / sleep quality (binary) / take medicine to sleep (binary)*

*Notes: * Only for Usoc and HILDA*

***Exposure***:

Exposure 1=Employment status (reference: inactive)

***Adjustment***:

~~~
           1.b.a.: gender, age, age-square, education
           1.b.b.: + Marital status, presence of children, logged household incomes,
           caring responsibilities, professional occupation
           1.b.c.: + self-reported health, longstanding illness
~~~

**Stratification:**

~~~
           1.a.d.: Male (fully adjusted model (1.a.c.) only)
           1.a.e: Female (fully adjusted model (1.a.c.) only)
           1.a.f.: age-group 20-39 (fully adjusted model (1.a.c.) only)
           1.a.g: age-group 40-59 (fully adjusted model (1.a.c.) only)
           1.a.h: age-group 59-69 (fully adjusted model (1.a.c.) only)
           *Note: Remove gender/age from the control variables*
~~~

### MODEL 2

#### Model 2.a. Two-way fixed effects (including longitudinal weight)

***Outcomes***: Sleeping hours (numeric, log) / trouble falling asleep (binary) / lost sleep over worry (binary) ^#^ / sleep quality (binary) / take medicine to sleep (binary)*

*Notes:* ^#^ *Not for HILDA, * Only for Usoc and HILDA*

***Exposure***:

Exposure 2=Working time (reference: not in employment)

***Adjustment***:

~~~
           2.a.a.: none
           2.a.b.: + Marital status, presence of children, logged household incomes,
           caring responsibilities, professional occupation
           2.a.c.: + self-reported health, longstanding illness
~~~

**Stratification:**

~~~
           2.a.d.: Male (fully adjusted model (2.a.c.) only)
           2.a.e.: Female (fully adjusted model (2.a.c.) only)
           2.a.f.: age-group 20-39 (fully adjusted model (2.a.c.) only)
           2.a.g: age-group 40-59 (fully adjusted model (2.a.c.) only)
           2.a.h: age-group 59-69 (fully adjusted model (2.a.c.) only)
           2a.i.: Non-professional occupations (fully adjusted model (2.a.c.) only)
           2a.j.: Non-professional occupations (fully adjusted model (2.a.c.) only)
~~~

#### Model 2.b. Mixed effect (including longitudinal weight)

***Outcomes***: Sleeping hours (numeric, log) / trouble falling asleep (binary) / lost sleep over worry (binary) ^#^ / sleep quality (binary) / take medicine to sleep (binary)*

*Notes:* ^#^ *Not for HILDA, * Only for Usoc and HILDA*

***Exposure***:

Exposure 2= Working time (reference: not in employment)

***Adjustment***:

~~~
           2.b.a.: gender, age, age-square, education
           2.b.b.: + Marital status, presence of children, logged household incomes, aring
           responsibilities, professional occupation
           2.b.c.: + self-reported health, longstanding illness
~~~

**Stratification:**

~~~
           2.a.d.: Male (fully adjusted model (2.a.c.) only)
           2.a.e.: Female (fully adjusted model (2.a.c.) only)
           2.a.f.: age-group 20-39 (fully adjusted model (2.a.c.) only)
           2.a.g: age-group 40-59 (fully adjusted model (2.a.c.) only)
           2.a.h: age-group 59-69 (fully adjusted model (2.a.c.) only)
           2a.i.: Non-professional occupations (fully adjusted model (2.a.c.) only)
           2a.j.: Non-professional occupations (fully adjusted model (2.a.c.) only)
           *Note: Remove gender/age from the control variables*
~~~

### MODEL 3

#### Model 3.a. Two-way fixed effects (including longitudinal weight)

***Outcomes***: Sleeping hours (numeric, log) / trouble falling asleep (binary) / lost sleep over worry (binary) ^#^/ sleep quality (binary) / take medicine to sleep (binary)*

*Notes:* ^#^ *Not for HILDA, * Only for Usoc and HILDA*

***Exposure***:

Exposure 3=interaction, Working time (reference: not in employment) * job security (reference: permanent)

***Adjustment***:

~~~
           3.a.a.: none
           3.a.b.: + Marital status, presence of children, logged household incomes,
           caring responsibilities, professional occupation
           3.a.c.: + self-reported health, longstanding illness
~~~

**Stratification:**

~~~
           3.a.d.: Male (fully adjusted model (3.a.c.) only)
           3.a.e.: Female (fully adjusted model (3.a.c.) only)
           3.a.f.: age-group 20-39 (fully adjusted model (3.a.c.) only)
           3.a.g: age-group 40-59 (fully adjusted model (3.a.c.) only)
           3.a.h: age-group 59-69 (fully adjusted model (3.a.c.) only)
           3.a.i.: Non-professional occupations (fully adjusted model (3.a.c.) only)
           3.a.j.: Non-professional occupations (fully adjusted model (3.a.c.) only)
~~~

#### Model 3.b. Mixed effect (including longitudinal weight)

***Outcomes***: Sleeping hours (numeric, log) / trouble falling asleep (binary) / lost sleep over worry (binary) ^#^ / sleep quality (binary) / take medicine to sleep (binary)*

*Notes:* ^#^ *Not for HILDA, * Only for Usoc and HILDA*

***Exposure***:

Exposure 3=interaction, Working time (reference: not in employment) * job security (reference: permanent)

***Adjustment***:

~~~
           3.b.a.: gender, age, age-square, education
           3.b.b.: + Marital status, presence of children, logged household incomes, aring responsibilities, professional occupation
           3.b.c.: + self-reported health, longstanding illness
~~~

**Stratification:**

~~~
           3.a.d.: Male (fully adjusted model (3.a.c.) only)
           3.a.e.: Female (fully adjusted model (3.a.c.) only)
           3.a.f.: age-group 20-39 (fully adjusted model (3.a.c.) only)
           3.a.g: age-group 40-59 (fully adjusted model (3.a.c.) only)
           3.a.h: age-group 59-69 (fully adjusted model (3.a.c.) only)
           3.a.i.: Non-professional occupations (fully adjusted model (3.a.c.) only)
           3.a.j.: Non-professional occupations (fully adjusted model (3.a.c.) only)
           *Note: Remove gender/age from the control variables*
~~~

Note: Provide the **coefficients when linear and OR when binary, significance and 95%CI**.

Calculate **Hausman test** for fully adjusted model (c) and each outcome:

|  | **Outcome** | **USoc** | **JHPS** | **SOEP** | **HILDA** |
| --- | --- | --- | --- | --- | --- |
| **1.a.c. / 1.b.c.** | Sleeping hours |  |  |  |  |
|  | Trouble falling asleep |  |  |  |  |
|  | Lost sleep over worry |  |  |  | × |
|  | Sleep quality |  |  |  |  |
| **2.a.c. / 2.b.c.** | Sleeping hours |  |  |  |  |
|  | Trouble falling asleep |  |  |  |  |
|  | Lost sleep over worry |  |  |  | × |
|  | Sleep quality |  |  |  |  |
| **3.a.c. / 3.b.c.** | Sleeping hours |  |  |  |  |
|  | Trouble falling asleep |  |  |  |  |
|  | Lost sleep over worry |  |  |  | × |
|  | Sleep quality |  |  |  |  |

### Sensitivity analyses

1. Replicate all the analyses for JHPS using the average sleeping time combining weekend and weekdays where: Average sleep time= (weekdays sleep *5 + weekend sleep *2)/7
2. Replicate all the fully adjusted analyses for four datasets using the last modality (4) only to generate binary variables for trouble falling asleep, lost sleep over worry and sleep quality instead of modalities 3-4 as in the main analyses.

## Discussion

Japan and Australia are characterized by high working hours that are not equally distributed among the working population and a labour market largely stratified by gender. By contrast, average working hours are lower in the United Kingdom and Germany. In addition, the gender distribution in employment among the four countries is also uneven. Similarly, sleep duration is higher in the other three countries than it is in Japan, with sleeping time increasing in the former and decreasing in the latter over the past decades. While the relationships between employment and working hours, on the one hand, and sleep duration and quality, on the other hand, are well established at the country level, few studies employ a global cross-national perspective to address how different employment contexts affect sleep. The present study aims to address this question using parallel analyses on four distinct longitudinal surveys. This is a novel approach to this topic, and we expect to answer the following questions: (1) Are some specific employment statuses associated with lower sleep duration or poor sleep quality and are these associations similar among Japan, the UK, Germany, and Australia?; (2) Are non-standard forms of employment including non-permanent contract and contract work associated with poorer sleep quality and to what extent?; (3) Is there a gender divide in sleep duration and quality and does employment and working time patterns across genders explain country differences in such a divide?; (4) Do employed workers trade off employment duration for sleep in the same way among Japan, the UK, Germany, and Australia?.

## Data Availability

This study uses four datasets that are available upon request.
Access to the Japan Household Panel Survey micro-data is available upon request via the Keio University (Japan) research portal: https://www.pdrc.keio.ac.jp/en/paneldata/datasets/jhpskhps/
Access to Understanding Society (UKHLS) micro-data is available upon request via the UK data archive: https://www.data-archive.ac.uk/
Access to German Socio-Economic Panel (SOEP) micro-data is available upon request via: https://www.eui.eu/Research/Library/ResearchGuides/Economics/Statistics/MicroDataSet
Access to the Household, Income and Labour Dynamics (HILDA) micro-data is available upon request via: https://dataverse.ada.edu.au/dataverse/DSSLongitudinalStudies

