## Supplementary files for "Cross-national comparison of the relationship between working hours and employment status and sleep duration and quality among Australia, Germany, Japan and the United Kingdom. A research protocol"

**Supplementary file S1. Employment and sleep data availability in JHPS, UKHLS, SOEP and HILDA.**

| **Dataset** | **Cohort** | **Variables** |  |  |  | **Waves** | | | | | | | | | | | | | | | | | | |
| --- | --- | --- | --- | --- | --- | --- | --- | --- | --- | --- | --- | --- | --- | --- | --- | --- | --- | --- | --- | --- | --- | --- | --- | --- |
| **JHPS (Japan)** | JHPS |  |  |  |  |  |  |  |  |  | **2009** | **2010** | **2011** | **2012** | **2013** | **2014** | **2015** | **2016** | **2017** | **2018** | **2019** | **2020** | **2021** | **2022** |
|  |  | Sleeping hours (weekdays) |  |  |  |  |  |  |  |  |  |  | X | X | X | X | X | X | X | X | X | X | X | X |
|  |  | Sleeping hours (weekend) |  |  |  |  |  |  |  |  |  |  | X | X | X | X | X | X | X | X | X | X | X | X |
|  |  | Trouble falling asleep |  |  |  |  |  |  |  |  | X | X | X | X | X |  |  |  |  |  |  |  |  |  |
|  |  | Lost much sleep over worry |  |  |  |  |  |  |  |  |  |  |  |  |  | X | X | X | X | X | X |  |  |  |
|  |  | Sleep quality |  |  |  |  |  |  |  |  |  |  |  |  |  |  |  |  |  |  |  |  | X | X |
|  |  | Working time (incl overtime) |  |  |  |  |  |  |  |  | X | X | X | X | X | X | X | X | X | X | X | X | X | X |
|  |  | Employment Status |  |  |  |  |  |  |  |  | X | X | X | X | X | X | X | X | X | X | X | X | X | X |
|  |  | Job satisfaction |  |  |  |  |  |  |  |  |  |  |  |  |  | X | X | X | X | X | X | X | X | X |
|  | KHPS |  |  |  |  | **2004** | **2005** | **2006** | **2007** | **2008** | **2009** | **2010** | **2011** | **2012** | **2013** | **2014** | **2015** | **2016** | **2017** | **2018** | **2019** | **2020** | **2021** | **2022** |
|  |  | Sleeping hours (weekdays) |  |  |  |  | X | X |  | X | X | X | X | X | X | X | X | X | X | X | X | X | X | X |
|  |  | Sleeping hours (weekend) |  |  |  |  |  |  |  |  |  |  |  |  |  | X | X | X | X | X | X | X | X | X |
|  |  | Trouble falling asleep |  |  |  |  | X | X |  | X | X | X | X | X | X |  |  |  |  |  |  |  |  |  |
|  |  | Lost much sleep over worry |  |  |  |  |  |  |  |  |  |  |  |  |  | X | X | X | X | X | X |  |  |  |
|  |  | Sleep quality |  |  |  |  |  |  |  |  |  |  |  |  |  |  |  |  |  |  |  |  | X | X |
|  |  | Working time (incl overtime) |  |  |  | X | X | X | X | X | X |  | X | X | X | X | X | X | X | X | x | X | X | X |
|  |  | Employment Status |  |  |  | X | X | X | X | X | X | X | X | X | X | X | X | X | X | X | X | X | X | X |
|  | KHPS+JHPS |  |  |  |  | **2004** | **2005** | **2006** | **2007** | **2008** | **2009** | **2010** | **2011** | **2012** | **2013** | **2014** | **2015** | **2016** | **2017** | **2018** | **2019** | **2020** | **2021** | **2022** |
|  |  | Sleeping hours (weekdays) |  |  |  |  | X | X |  | X | X | X | X | X | X | X | X | X | X | X | X | X | X | X |
|  |  | Sleeping hours (weekend) |  |  |  |  |  |  |  |  |  |  | X | X | X | X | X | X | X | X | X | X | X | X |
|  |  | Trouble falling asleep |  |  |  |  | X | X |  | X | X | X | X | X | X |  |  |  |  |  |  |  |  |  |
|  |  | Lost much sleep over worry |  |  |  |  |  |  |  |  |  |  |  |  |  | X | X | X | X | X | X |  |  |  |
|  |  | Sleep quality |  |  |  |  |  |  |  |  |  |  |  |  |  |  |  |  |  |  |  |  | X | X |
|  |  | Working time (incl overtime) |  |  |  | X | X | X | X | X | X | X | X | X | X | X | X | X | X | X | X | X | X | X |
|  |  | Employment Status |  |  |  | X | X | X | X | X | X | X | X | X | X | X | X | X | X | X | X | X | X | X |
| **Usoc (UK)** | UKHLS |  |  |  |  |  |  |  |  |  | **2009** | **2010** | **2011** | **2012** | **2013** | **2014** | **2015** | **2016** | **2017** | **2018** | **2019** | **2020** | **2021** | **2022** |
|  |  | Sleeping hours (weekdays) |  |  |  |  |  |  |  |  | X |  |  | X |  |  | X |  |  | X |  |  | X |  |
|  |  | Sleeping hours (weekend) |  |  |  |  |  |  |  |  |  |  |  |  |  |  |  |  |  |  |  |  |  |  |
|  |  | Trouble falling asleep |  |  |  |  |  |  |  |  |  |  |  | X |  |  | X |  |  | X |  |  | X |  |
|  |  | Lost much sleep over worry |  |  |  |  |  |  |  |  | X | X | X | X | X | X | X | X | X | X | X | X | X | X |
|  |  | Sleep quality |  |  |  |  |  |  |  |  |  |  |  | X |  |  | X |  |  | X |  |  | X |  |
|  |  | Take medicine to sleep |  |  |  |  |  |  |  |  |  |  |  | X |  |  | X |  |  | X |  |  | X |  |
|  |  | Working time (incl overtime) |  |  |  |  |  |  |  |  | X | X | X | X | X | X | X | X | X | X | X | X | X | X |
|  |  | Employment Status |  |  |  |  |  |  |  |  | X | X | X | X | X | X | X | X | X | X | X | X | X | X |
| **Usoc (UK)** | SOEP |  |  |  |  |  |  | **2006** | **2007** | **2008** | **2009** | **2010** | **2011** | **2012** | **2013** | **2014** | **2015** | **2016** | **2017** | **2018** | **2019** | **2020** | **2021** |  |
|  |  | Sleeping hours (weekdays) |  |  |  |  |  |  |  | X | X | X | X | X | X |  | X |  | X |  | X | X | X |  |
|  |  | Sleeping hours (weekend) |  |  |  |  |  |  |  |  |  |  |  |  |  |  |  |  |  |  |  |  |  |  |
|  |  | Trouble falling asleep |  |  |  |  |  | X |  |  |  |  | X | X |  |  |  | X |  |  |  |  | X |  |
|  |  | Lost much sleep over worry |  |  |  |  |  |  |  |  |  |  | X |  | X |  | X |  | X |  | X |  | X |  |
|  |  | Sleep quality |  |  |  |  |  |  |  | X | X | X | X | X | X | X | X | X | X | X | X | X | X |  |
|  |  | Working time |  |  |  |  |  | X | X | X | X | X | X | X | X | X | X | X | X | X | X | X | X | X |
|  |  | Employment Status |  |  |  |  |  | X | X | X | X | X | X | X | X | X | X | X | X | X | X | X | X | X |
| **Usoc (UK)** | HILDA |  | **2001** | **2002** | **2003** | **2004** | **2005** | **2006** | **2007** | **2008** | **2009** | **2010** | **2011** | **2012** | **2013** | **2014** | **2015** | **2016** | **2017** | **2018** | **2019** | **2020** | **2021** | **2022** |
|  |  | Sleeping hours (weekdays) |  |  |  |  |  |  |  |  |  |  |  |  | X |  |  |  | X |  |  |  | X |  |
|  |  | Sleeping hours (weekend) |  |  |  |  |  |  |  |  |  |  |  |  |  |  |  |  |  |  |  |  |  |  |
|  |  | Trouble falling asleep |  |  |  |  |  |  |  |  |  |  |  |  |  |  |  |  |  |  |  |  |  |  |
|  |  | Sleep quality |  |  |  |  |  |  |  |  |  |  |  |  |  |  |  |  |  |  |  |  |  |  |
|  |  | Take medicine to sleep |  |  |  |  |  |  |  |  |  |  |  |  |  |  |  |  |  |  |  |  |  |  |
|  |  | Working time | X | X | X | X | X | X | X | X | X | X | X | X | X | X | X | X | X | X | X | X | X | X |
|  |  | Employment Status | X | X | X | X | X | X | X | X | X | X | X | X | X | X | X | X | X | X | X | X | X | X |

**Supplementary file S2. Main variables harmonization across JHPS, UKHLS, SOEP and HILDA**

|  | **JHPS** | | |
| --- | --- | --- | --- |
|  | Question | Modalities | *Waves* |
| **Sleeping hours (weekdays)** | Please, write your usual sleeping hours on weekdays | Numeric, in hours | *2009 to 2022* |
| **Sleeping hours (weekend)** | Please, write your usual sleeping hours on weekends | Numeric, in hours | *2011 to 2022* |
| **Trouble falling asleep** | I have trouble getting to sleep | 1 (often) - 4 (never) | *2009 to 2013* |
| **Lost much sleep over worry** | Lost much sleep over worry? | 1 (never) - 4 (often) | *2014 to 2019* |
| **Sleep quality** | How would you rate the overall quality of your sleep over the past month? | 1 (excellent) - 4 (very bad) | *2021-2022* |
| **Working time (incl overtime)** | Hours worked per week including over time | Numeric, in hours | *All waves* |
| **Employment Status (1)** | Main activity | Employed full-time, employed part-time, contract worker, self-employed, unemployed, retired, other inactive | *All waves* |
| **Employment Status (2)** |  |  |  |
| **Age** | Age | Year of birth | *All waves* |
| **Gender** | Gender | Male, female | *All waves* |
| **Marital Status** | Marital Status | Married, Single, don't know | *All waves* |
| **Self-reported health** | How would you rate your health on the whole? | (1) Good - (5) Bad | *All waves* |
| **Longstanding illness** | Were you hospitalized or did you go to a hospital for treatment of a disease or injury in the last year? | (1) Hospitalizd, (2) visited hospital, (3) both, (4) Neither | *All waves* |
| **Highest level of education (degree / no degree)** | Choose a school that you last attended. | (1) Junior high school, (2) high school, (3) Junior high school, (4) four-year university, (5) graduate school, (6) other | *All waves* |
| **Presence of children** | Number of children |  |  |
| **Caring responsibilities** | Does any member of your family need nursing care? | (1) Yes (in nursing home); (2) Yes (living together); (3) other; (4) no | *All waves* |
| **Commuting time** | NA | NA | *NA* |
| **Professional / non-professional** | Employment position | Full-tile employee with no title, full-time regular employee with title, full-time regular employee manager | *All waves* |
|  | **Usoc** | | |
|  | Question | Modalities | *Waves* |
| **Sleeping hours (weekdays)** | Average sleeping hours per day (no distinction between weekdays and weekand) | Numeric, in hours | *2009, 2012, 2015, 2018, 2021* |
| **Sleeping hours (weekend)** |  |  |  |
| **Trouble falling asleep** | cannot get to sleep within 30 mins | (1) not during the past month - (4) more than once most nights | *2012, 2015, 2018, 2021* |
| **Lost much sleep over worry** | (As part of GHQ) Loss of sleep | 1 (not at all) - 4 (much more than usual) | *2009 to 2021* |
| **Sleep quality** | During the past month, how would you rate your sleep quality overall? | 1 (very good) - 4 (very bad) | *2012, 2015, 2018, 2021* |
| **Working time (incl overtime)** | Hours worked per week including over time | Numeric, in hours | *All waves* |
| **Employment Status (1)** | Main activity | Employed, slef-employed, unemployed, inactive | *All waves* |
| **Employment Status (2)** | Is your current job is permanent or temporary? (Leaving aside your own personal intentions and circumstances) **JBTERM1** | (1) Permanent, (2) Temporary in some way |  |
| **Age** | Age | Year of birth | *All waves* |
| **Gender** | Gender | Male, female | *All waves* |
| **Marital Status** | Marital Status | Married, non-married | *All waves* |
| **Self-reported health** | How would you rate your health on the whole? | (1) Excellent - (5) Bad | *All waves* |
| **Longstanding illness** |  |  |  |
| **Highest level of education (degree / no degree)** |  |  |  |
| **Presence of children** |  |  |  |
| **Caring responsibilities** |  |  |  |
| **Commuting time** |  |  |  |
| **Professional / non-professional** | NS-SEC | Large employers & higher management; Higher professional; Lower management & professional; Intermediate; Small employers & own account; Lower supervisory & technical; Semi-routine; Routine | *All waves* |
|  | **SOEP** |  |  |
|  | Question | Modalities | *Waves* |
| **Sleeping hours (weekdays)** | How many hours do you sleep on average on a normal day during the working week? | Numeric, in hours | *2008-2013,2015,2017,2019-2021* |
| **Sleeping hours (weekend)** | How many hours on a normal weekend day? |  |  |
| **Trouble falling asleep** | Has a doctor ever diagnosed you with sleep disorder? | yes | *2006, 2011, 2012, 2016, 2021* |
| **Lost much sleep over worry** | If I put off something that needs to be done that day, I can't sleep at night | 1 (strongly disagree) - 4 (strongly agree) | *2011, 2013, 2015, 2017, 2019, 2021* |
| **Sleep quality** | How satisfied are you with sleep? | (0)completely dissatisfied-(10)completely satisfied | *2008-2021* |
| **Working time (incl overtime)** | Hours worked per week | Numeric, in hours | *All waves* |
| **Employment Status (1)** | Employment Status of Individual | Full-Time Employment Part-Time Employment Occasion employment | *All waves* |
| **Employment Status (2)** |  |  |  |
| **Age** | Age | Year of birth | *All waves* |
| **Gender** | Gender | Male, female | *All waves* |
| **Marital Status** | Marital Status | Married, non-married | *All waves* |
| **Self-reported health** | How satisfied are you with your current health? | (0)completely dissatisfied-(10)completely satisfied | *All waves* |
| **Longstanding illness** | Do you have any health problems that limit your normal daily activities? | (1)Yes, severely  (2)Yes, somewhat  (3) No, not at all | *2001-2021* |
| **Highest level of education (degree / no degree)** | The highest level of education | Less than High School High School More than High School | *All waves* |
| **Presence of children** | Number of children in household | Numeric | All waves |
| **Caring responsibilities** | How many hours do you spend on childcare on a typical weekday, Saturday, and Sunday? | Numeric | *2009-2021* |
| **Commuting time** |  |  |  |
| **Professional / non-professional** | What is your current occupational status | Self-employed (including family members working for the self-employed)  Blue-collar worker (also in agriculture)  Civil servant (including judges and professional soldiers)  Apprentice / trainee / intern  White-collar worker | *All waves* |
|  | **HILDA** |  |  |
|  | Question | Modalities | *Waves* |
| **Sleeping hours (weekdays)** | How many hours of actual sleep do you usually get on a workday night (currently employed) | Numeric, in hours | *2013, 2017, 2021* |
| **Sleeping hours (weekend)** | Hours of sleep per week |  |  |
| **Trouble falling asleep** | *Had trouble sleeping because cannot get to sleep within 30 minutes;Had trouble sleeping because cough or snore loudly; Had trouble staying awake while driving, eating meals or engaging in social activity; Had taken medicine to help sleep; Had trouble sleeping because wake up in the middle of the night or early in the morning* | *(1) Not during the past month (2) Less than once a week (3) Once or twice a week (4) Three or four times a week (5) Five or more times a week* | *2013, 2017, 2021* |
| **Lost much sleep over worry** |  |  |  |
| **Sleep quality** | In the past month, how would you rate your sleep overall | *(1)Very good (4) Very bad* | *2013, 2017, 2021* |
| **Working time (incl overtime)** | Hours per week usually worked in all jobs | Numeric, in hours | *All waves* |
| **Employment Status (1)** | Employment status | Employed - usually works 35+ hours per week; Employed - usually works less than 35 hours per week; Not employed but is looking for work; Retired; Home duties; Non-working student; Other | *All waves* |
| **Employment Status (2)** |  |  |  |
| **Age** | Age | Year of birth | *All waves* |
| **Gender** | Gender | Male, female | *All waves* |
| **Marital Status** | Marital Status | Married, non-married | *All waves* |
| **Self-reported health** | Self-assessed health | (1) Excellent (5) poor | *All waves* |
| **Longstanding illness** | Long term health condition | Yes No | *All waves* |
| **Highest level of education (degree / no degree)** | Highest education level achieved | 1Postgrad - masters or doctorate 2 Grad diploma, grad certificate 3 Bachelor or honours 4 Adv diploma, diploma 5 Cert III or IV 8 Year 12 9 Year 11 and below | *All waves* |
| **Presence of children** | Number of dependent children | Numeric | All waves |
| **Caring responsibilities** | Annual child care total cost | Numeric | *2003-2022* |
| **Commuting time** |  |  |  |
| **Professional / non-professional** | ISCO-88 2-digit, Occupation current main job | Professional if jbm682>1 & jbm682<30 Non-professional if jbm682>24 | *All waves* |
|  | **Harmonization** | **Sensitivity checks** |  |
| **Sleeping hours (weekdays)** | Logged Numeric | Average sleeping time (mean of weekdays and weekend sleep for JHPS) |  |
| **Sleeping hours (weekend)** |  |  |  |
| **Trouble falling asleep** | 0 (1, 2); 1 (3, 4) | 0 (1,2,3); 1 (4) |  |
| **Lost much sleep over worry** | 0 (1, 2); 1 (3, 4) | 0 (1,2,3); 1 (4) |  |
| **Sleep quality** | 0 (1, 2); 1 (3, 4) | 0 (1,2,3); 1 (4) |  |
| **Working time (incl overtime)** | (1) 1 to 20 hours/week; (2) 21-34 hours/week; (3) 35-48 hours/week; (4) 48+ hours/week; (5) not in employment (reference) |  |  |
| **Employment Status (1)** | (1) permanent employment (part-time and full-time), (2) temporary employment or contract work, (3) self-employment, (4) unemployment, (5) inactivity (reference category). |  |  |
| **Employment Status (2)** |  |  |  |
| **Age** | Age (in years of age) | Age-square |  |
| **Gender** | (0) Male (Ref.), (1) Female |  |  |
| **Marital Status** | (0) Married (Ref.), (1) non-married |  |  |
| **Self-reported health** | (1) Excellent (reference) - (5) Bad |  |  |
| **Longstanding illness** | (1)Yes (0)No |  |  |
| **Highest level of education (degree / no degree)** | University degree (0), No university degree (1) |  |  |
| **Presence of children** |  |  |  |
| **Caring responsibilities** |  |  |  |
| **Commuting time** | Data not available in JHPS | One extra layer of adjustment for Usoc |  |
| **Professional / non-professional** | (1)Pro (0)Non-pro |  |  |
